# TOWARDS AN AI-DRIVEN REGISTRY FOR POSTOPERATIVE COMPLICATIONS: A PROOF-OF-CONCEPT STUDY EVALUATING THE OPPORTUNITIES AND CHALLENGES OF AI-MODELS

**DOI:** 10.1101/2025.04.07.25325369

**Authors:** Emilie E. Dencker, Andreas Skov Millarch, Alexander Bonde, Anders Troelsen, Jens Winther Jensen, Martin Sillesen

**Affiliations:** Dep. Of Organ Surgery and Transplantation, Copenhagen University Hospital, Rigshospitalet, Denmark; Center for Surgical Translational and Artificial intelligence Research (CSTAR), Copenhagen University Hospital, Rigshospitalet, Denmark; Aiomic Aps, Denmark; Dep. of Orthopedics, Copenhagen University Hospital, Hvidovre, Denmark; Institute of Clinical Medicine, University of Copenhagen Medical School; The Danish Healthcare Quality Institute (DHQI)

## Abstract

**Background:** Continuous quality improvement is essential in surgery, with clinical registries and quality improvement programs (QIPs) playing a key role. Postoperative complications (PCs) require substantial resources to manage, yet traditional QIPs are expensive and often lays a significant labor burden on clinicians in data collection. Artificial intelligence (AI), particularly natural language processing (NLP), offers a potential solution by automating and streamlining these processes, but models can be optimized for optimal sensitivity or positive predictive value. This study aimed to develop a mock-up automated registry for PCs using NLP algorithms and evaluate the effects of optimization strategies for surgical quality control. We hypothesized using NLP to obtain longitudinal overviews of key quality metrics is feasible, but that optimization strategies impacted on the observed rate of PCs and thus how quality management and surveillance would be affected in a real-world setting.

**Methods:** We analyzed 100,505 surgical cases from 12 Danish hospitals between 2016 and 2022. Previously validated NLP models were applied to detect seven types of PCs, using two different threshold settings: a set of thresholds optimized for positive predictive value (PPV or Precision), referred to as F-score of 0.5, and a set of thresholds optimized for sensitivity, referred to as F-score of 2. Trends in PC rates over time were assessed, and hospital-level variations were examined using logistic regression models adjusted for age, sex, and comorbidity.

**Results:** The NLP models detected 8,512 or 15,892 PCs, depending on threshold selection, corresponding to total PC rates of 9.14% and 17.1%, respectively. Most PCs showed stable or increasing trends over time, regardless of threshold setting. Hospital-level analyses similarly revealed stable or rising PC rates in most institutions. Regression analyses demonstrated that threshold selection significantly influenced findings, impacting hospital comparisons.

**Conclusion:** This study demonstrates that NLP can be used for automated PC detection in surgical quality monitoring. However, threshold selection and additional performance metrics, such as precision-recall curves (PPV-Sensitivity curves), must be carefully considered to ensure reliable and meaningful results beyond traditional Receiver Operator Area Under the Curve (ROC AUC) evaluation.

## INTRODUCTION

According to the World Health Organization (WHO), adverse events are among the top ten leading causes of death and disability[1]. The Salzburg Statement on Global Principles for Measuring Patient Safety further emphasizes that this ranking will not improve without a coordinated, global effort that is both effective and efficient[1]. Therefore, continuous quality improvement, refinement of skills and optimized patient outcomes, are all fundamental components in modern surgery. As a result, clinical registries and quality improvement programs serve as a cornerstone for understanding and improving outcomes[2,3]. Evidence from the literature demonstrates that participating in clinical registries and quality improvement programs, such as the American College of Surgeons National Quality Improvement Program (ACS NSQIP), lead to improved outcomes, reduced adverse events, and lower mortality rates. Furthermore, these programs have been proven to be cost-effective despite their relatively high expenses[2–4].

Conversely, the literature also highlights the significant number of resources (physical and financial) spent on managing adverse event, such as postoperative complications (PCs). PCs are associated with prolonged admissions, increased need for ICU care, reoperations, and readmissions, all of which place considerable strain on healthcare systems[5–8]. Adding to this strain, healthcare systems worldwide face unprecedented challenges as demographic shifts have led to growing patient populations[9], while staff shortages have further increased the pressure on healthcare resources. Despite this well-established evidence, many countries lack comprehensive national registries that cover all surgical specialties and relevant outcomes. Instead, existing registries, are often limited to specific surgical specialties or even individual procedures[10].

In the context of the emerging AI era, where automation is increasingly possible, utilizing machine learning for surgical quality improvement presents a clear opportunity. In recent years, natural language processing (NLP) has demonstrated its ability to detect and extract various details from electronic healthcare records (EHRs) including postoperative complications[11–15]. Automating this traditionally manual process could free up critical human resources for more essential tasks while enabling real-time surveillance of PCs. Additionally, this real-time surveillance could enable timely and relevant interventions to reduce PC incidence. This approach aligns with key principles of the Salzburg Statement on Measuring Patient Safety by facilitating real-time data collection and analysis while reducing the burden of data collection[1].

Moreover, a national – or potentially international – automated registry for postoperative complications, has the potential to standardize surgical quality across hospitals or even across national borders, thereby promoting greater equity in healthcare systems.

This study aimed to develop a mock-up automated national registry for postoperative complications in a Danish setting while exploring the advantages and challenges of using NLP algorithms in surgical quality improvement. Using a suite of previously validated NLP models for PC capture[11,12,16], we hypothesized that existing models could successfully create an automated PC registry and reveal variations in surgical quality across the included hospitals, but also that optimizing models for either positive predictive value or sensitivity would result in discrepant incidences of PCs captured and thus directly impact on perceived level of surgical quality.

## METHODS

### Approvals

The study was approved by the Institutional Review Board (IRB) overseeing retrospective patient studies in Denmark, the Danish Patient Safety Board (Styrelsen for Patientsikkerhed, approval #31-1521-182), and the Danish Capital Region Data Safety Board (Videnscenter for Dataanmeldelser, approval #P-2020-180). Since the study was retrospective, used deidentified data, and had no direct patient contact, Danish law did not require informed consent.

### Source of data

The study data was derived from a dataset encompassing all patients who underwent surgical procedures at one of 12 hospitals in Denmark’s Capital Region and the Zealand region over a five-year period (January 1^st^, 2017, and December 31^st^, 2021). Combined, these regions have a population of 2.7 million, with the included hospitals responsible for providing all healthcare services free of charge within this population.

The dataset contained electronic healthcare record (EHR) data, including clinical notes (e.g., admission, procedure, progress, and discharge notes), biochemical values (e.g., sodium, white blood cells count, hematocrit, albumin) and vital signs (e.g., temperature, blood pressure, pulse). Additionally, it included demographic details, ICD-10 diagnosis codes for acute conditions, comorbidities, postoperative complications, and hospital location. For each patient, data collection began on the day of the primary surgical procedure and continued for 30 days post-operatively. The dataset encompassed all surgical specialties and included both elective and acute procedures

Since the study aimed to develop a mock-up automated national registry for postoperative complications (PCs) and assess potential variations in surgical quality across hospitals, it was essential to create a uniform subset of data. This ensured that any observed differences in postoperative complication rates were not simply due to variations in case complexity or patient health.

To achieve this, we selected the 10 most common surgical procedures that met the following criteria:

- *Performed at all participating hospitals*
- *Required general or regional (spinal/epidural) anesthesia*

Ophthalmologic and endoscopic procedures were excluded, as they typically carry lower complication risks and are often performed in outpatient settings.

The 10 included procedures spanned four of the largest surgical specialties: general surgery (laparoscopic cholecystectomy, laparoscopic appendectomy, laparoscopy (with biopsy), and laparoscopic inguinal hernia repair), orthopedic surgery (primary total hip arthroplasty, primary total knee arthroplasty and internal fixation of the distal radius), gynecologic and obstetric surgery (elective cesarean section and emergency cesarean section), and urologic surgery (transurethral resection of the bladder (TUR-B)).

Cases with missing values for any key variable (Case ID, Patient ID, age, sex, surgery start and end time, or planned procedure code) were excluded during data preprocessing.

### Postoperative complications

This study included seven early postoperative complications, defined as those occurring within 30 days postoperatively. The selected complications were urinary tract infection (UTI), pneumonia, superficial surgical site infection (SSSI), sepsis, septic shock, unplanned reoperation, and unplanned readmission. These complications were chosen because they are general rather than procedure-specific, making them broadly applicable across different surgical specialties, Additionally, they are associated with increased healthcare resource utilization, further emphasizing their clinical and economic impact[5–8]. Moreover, previous studies demonstrated that these seven complications yielded promising detection results when applying earlier developed natural language processing (NLP) models to electronic health records (EHRs)[11,12,16]. In those studies, manual data labeling for NLP model development followed the American College of Surgeons National Quality Improvement Program (ACS NSQIP) definitions of these complications which are available in Supplementary Material Table S1.

### Natural language processing (NLP) algorithms

The NLP models utilized in this study were previously developed to detect postoperative complications directly from EHRs. The models were trained and validated in prior studies within our research group, demonstrating high performance in extracting complications[11,12,16].

Briefly, model development was based on EHR data from hospitals in Denmark’s Capital and Zealand regions. Labeling of data was performed by a team of medical reviewers. One model was developed for each complication, using text vectorization with Term Frequency-Inverse Document Frequency (TF-IDF) and classification using Light Gradient Boosting Machine (LightGBM) algorithms.

Each model predicts the presence of a specific PC by outputting a probability score. Whether a case is classified as having a PC present, depends on the selected threshold, which is further explored in the following section.

Model performance was previously evaluated using Receiver Operator Area Under the Curve (ROC AUC), sensitivity, specificity, positive predictive value (PPV), and negative predictive value (NPV), with manual chart review serving as the gold standard. Across all included PCs, the models demonstrated high ROC AUC values (ranging from 0.890 to 0.998), sensitivity (0.701 to 0.955), specificity (0.964 to 0.997), PPV (0.244 to 0.947), and NPV (0.981 to 0.999) [11,12,16] Additionally, given the imbalance of the data, we assessed model performance using precision-recall (PR) curves, providing a more focused evaluation of precision and recall.

### Threshold selection

The NLP models used in this study are probabilistic classifiers, meaning they output a probability score rather than a simple binary yes/no classification for the presence of a PC. Therefore, a decision threshold is necessary to determine at what probability value a case is classified as positive for having a given PC. The choice of threshold directly impacts performance metrics including sensitivity, specificity, positive predictive value (PPV), and negative predictive value (NPV).

To illustrate how different threshold selections affect NLP model performance, we applied two distinct thresholding strategies by optimizing different F-scores. The F-score is a metric that balances PPV (precision) and sensitivity (recall), calculated as:

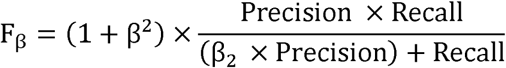

An F-score of 1 gives equal weight to PPV and sensitivity. Consequently, an F-score of 2 emphasizes sensitivity more than PPV, while an F-score of 0.5 prioritizes PPV over sensitivity. By selecting F2 and F0.5 for threshold optimization, we explicitly favor either sensitivity or precision, depending on the clinical setting.

1) **Precision-Optimized Threshold (F0.5-optimized)** This threshold was set to maximize the positive predictive value, thereby minimizing the number of false positives, which is beneficial in settings where overdiagnosis or unnecessary interventions is crucial.
2) **Sensitivity-Optimized Thresholds (F2-optimized)** This threshold was set to maximize sensitivity, thereby minimizing the number of false negatives, making this approach useful when the primary goal is to detect as many true cases as possible.

By assessing model outcomes by both threshold sets, we aimed to demonstrate how the same NLP models can yield different outcomes depending on the balance between sensitivity and PPV.

### Statistical analysis

Descriptive statistics were performed to summarize demographic characteristics and the number of PCs detected by the NLP models. Results are reported as whole numbers, percentages, and medians with interquartile ranges [Q1–Q3], where appropriate.

To illustrate the variability on PC detection based on threshold selection, results are presented using the two threshold settings: the PPV-Optimized Threshold (F0.5-optimized) and the Sensitivity-Optimized Threshold (F2-optimized), and the PC rates over time were reported as percentages – both total PC rates per hospital and individual rates per complication were performed.

As PC capture for national registries will be used to benchmark hospitals and surgical units, we deployed logistic regression models to assess associations between hospital location and individual PCs. Two separate models were developed for each PC – using both the PPV-Optimized Threshold (F0.5-optimized) and the Sensitivity-Optimized Threshold (F2-optimized). Each model included hospital location as the independent variables, adjusting for age, sex, and Elixhauser comorbidity score. Regression results are presented as odds ratios (ORs) with 95% confidence intervals (CIs) with corresponding p-value. For readability, full regression results are provided for one PC in the main text (superficial surgical site infection), while remaining regression results are available in the Supplementary Material, Table S2.

All statistical analyses were performed in Python version 3.10 using “statsmodels” version 0.14.4. P-valuesl<l0.05 was considered statistically significant.

## RESULTS

### Demographics

The initial dataset comprised 1,444,526 surgical cases, representing all patients who underwent surgical procedures at one of 12 Danish hospitals between January 1^st^ 2017 and December 31^st^ 2021. After applying inclusion criteria and removing cases with missing data, the final subset of data comprised 100,505 surgical cases corresponding to 93,095 unique patients. Of these, 62.2% were female, with a median age of 55 years (interquartile range [IQR]: 33-71) and a median BMI of 26.6 [23.5-30.7]. A complete demographic overview is provided in Table 1. Table 1 displays the included surgical specialties, procedures, and patient characteristics.

**Table 1.**
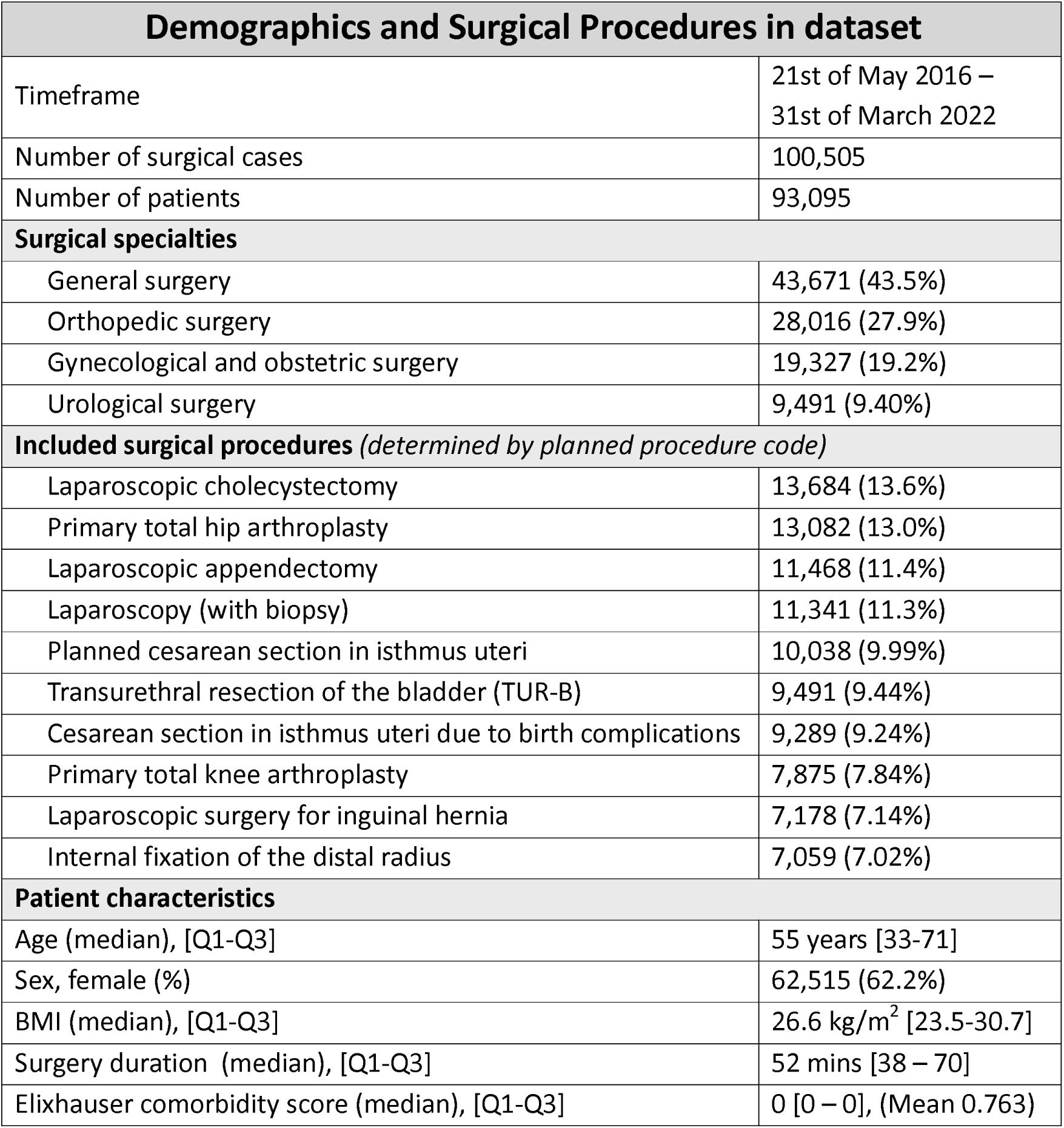
Demographics, surgical specialties, and included surgical procedures. Results are presented as percentages, medians with interquartile ranges [Q1-Q3], or means where appropriate.

### Postoperative complications in the “mock-up registry” - Trends over time

When applying the F0.5 thresholds, which prioritize positive predictive value, the models detected a total of 8,512 PCs within the dataset. This corresponds to a total PC rate of 9.14%. The distribution across the seven studied complications is detailed in Table 2. In contrast, applying the F2 thresholds, which prioritize sensitivity, resulted in the detection of 15,892 PCs within the dataset, corresponding to a total PC rate of 17.1%. The distribution of PCs under the F2 thresholds is also presented in Table 2.

**Table 2.** Number and percentage of postoperative complications detected using the F0.5 and F2 thresholds. Rates are calculated based on the number of patients.

| Postoperative Complication | F0.5 threshold applied | F2 threshold applied |
| --- | --- | --- |
| Urinary tract infection | 328 (0.352%) | 2,561 (2.75%) |
| Pneumonia | 389 (0.418%) | 1,052 (1.13%) |
| Sepsis | 309 (0.332%) | 771 (0.828%) |
| Septic shock | 47 (0.0505%) | 253 (0.272%) |
| Superficial surgical site infection | 1,118 (1.20%) | 2,229 (2.39%) |
| Unplanned reoperation | 1,911 (2.05%) | 3,783 (4.06%) |
| Unplanned readmission | 4,410 (4.74%) | 5,243 (5.63%) |
| Total PCs | 8,512 (9.14%) | 15,892 (17.1%) |

Thus, using the F2 thresholds nearly doubled the number of detected PCs compared to the F0.5 thresholds (15,892 vs. 8,512). This difference translates to an increase of eight percentage points in the total PC rate (9.14% vs. 17.1%) depending on the thresholds selection for the NLP models.

When examining the individual PC rates, we observed a clear difference in rates over time between the two sets of thresholds which are presented in Figure 1A-G. The trends over time for urinary tract infection (UTI), pneumonia, and SSSI remained relatively stable under both thresholds. For urinary tract infections (UTI), the F0.5 threshold resulted in a rate of 0.4% over time, whereas the F2 threshold produced a higher rate of 2.5-3.0%. Similarly, pneumonia rates were 0.4 % vs. 1.0% depending on the threshold used. For superficial surgical site infection (SSSI), the F0.5 threshold resulted in a rate of 1.0-1.2% over time, while the F2 threshold consistently yielded a higher rate of 2.2%.

**FIGURE 1A-G:**
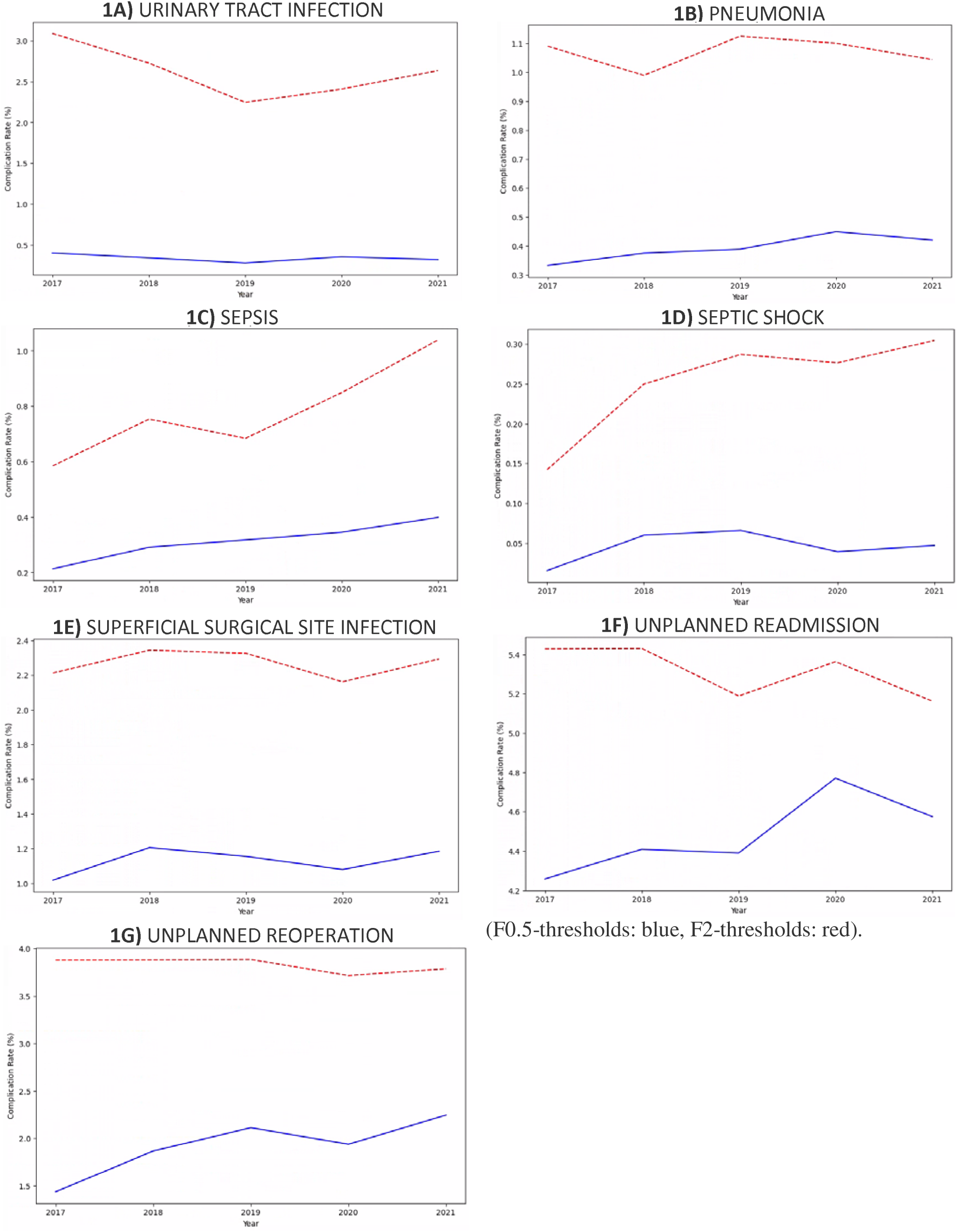
Individual PC rates over time

In contrast, sepsis, and septic shock both showed increasing trends over time regardless of the threshold applied. The F0.5 threshold detected sepsis rates increasing from 0.2% to 0.4%, whereas the F2 threshold showed a rise from 0.6 % to 1.1%. Septic shock followed a similar pattern, with rates increasing from 0.02% to 0.05% under F0.5 threshold, and from 0.15% to 0.30% under F2 threshold. Lastly, the trends for unplanned readmission and unplanned reoperation differed between the two thresholds. When applying the F0.5 thresholds, both PCs showed an increasing trend over time (unplanned reoperation from 1.5% to 2.25% and unplanned readmission from 4.3 % to 4.6%). However, when applying the F2 thresholds, the trend for unplanned reoperation remained stable at approximately 3.8%, while unplanned readmission showed a slight decline from 5.5% to 5.2%. All individual PC rates are displayed in Figure 1A-G.

Figure 2A and B illustrate the total PC rate over time for each hospital. Figure 2A presents the total PC rates using the F0.5 thresholds, while Figure 2B displays the rates using the F2 thresholds. In Figure 2A, the total PC rate in 2017 ranged between 3% (Hospital X) and 14% (Hospital Y). By 2021, this range had shifted to 8% (Hospital Z) to 14% (Hospital Y). Over the study period, eight hospitals exhibited increasing PC rates, while three hospitals showed a decline. One hospital started and ended the study period with the same PC rate of 14%, despite experiencing notable fluctuations in between (Hospital F).

**FIGURE 2A-B:**
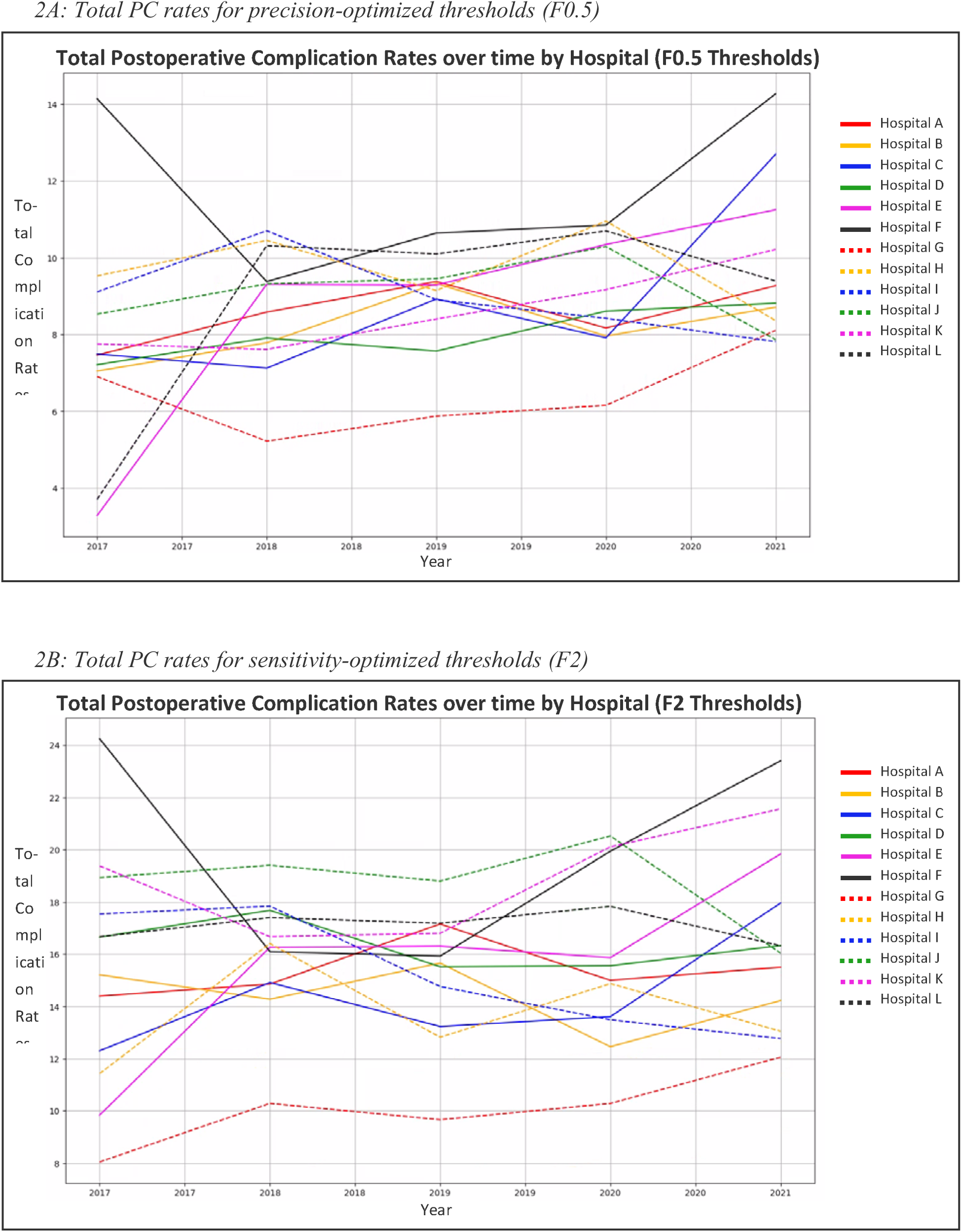
Total PC rates over time for each hospital for the different thresholds.

When applying the F2 thresholds (Figure 2B), the total PC rate in 2017 ranged from 8% (Hospital A) to 24% (Hospital B), while in 2021, it ranged from 12% (Hospital C) to 23% (Hospital D). Over time, six hospitals demonstrated an increasing trend, four hospitals showed a decrease, and two hospitals maintained stable PC rates throughout the study period.

### Precision - Recall curves

Figure 3 presents the precision-recall (PR) curves for all seven PCs, along with their respective Area Under the Precision-Recall Curve (AUC-PR) values. The AUC-PR scores range from 0.78 to 0.95, indicating strong overall model performance, though with some variation across different complications. The highest-performing models are those detecting SSSI, unplanned readmission, and unplanned reoperation, which maintain high precision across all recall levels, leading to fewer false positives. The sepsis and septic shock models follow with slightly more trade-off between precision and recall. In contrast, the UTI and pneumonia models demonstrate the lowest AUC-PR values, indicating that achieving high recall comes at the cost of lower precision, resulting in more false positives.

**Figure 3.**
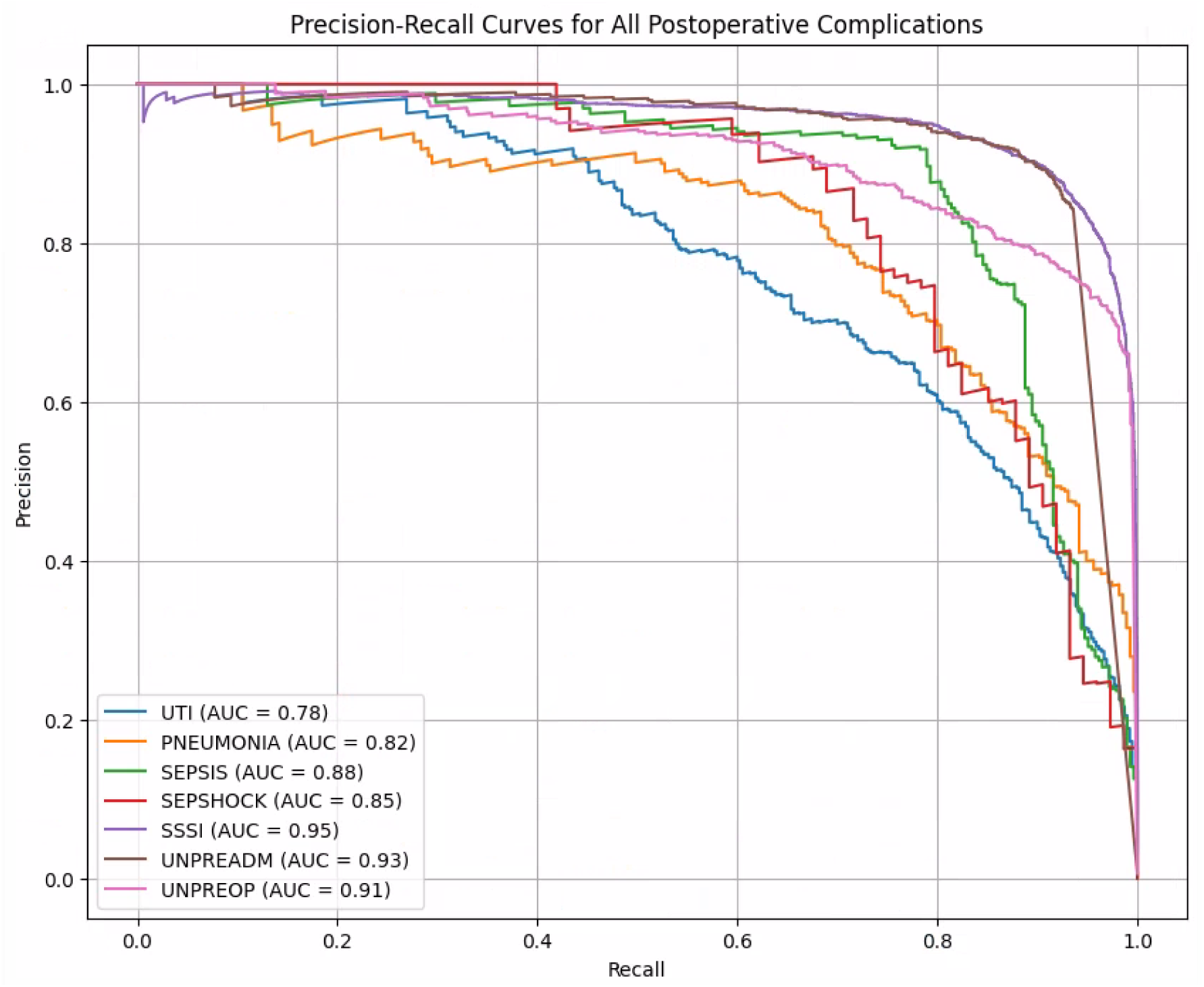
Precision-Recall curves for all studied postoperative complications including AUC-PR.

### Regression model results – does surgical quality vary across the included hospitals?

To assess whether the differences in total PC rates observed in Figure 2A and B were statistically significant, we developed multiple logistic regression models. Specifically, two models were created for each PC – one using the F0.5 thresholds and one using the F2 thresholds. The objective of these models was to determine whether any of the included hospitals were associated with a higher probability of a PC, after adjusting for age, sex (male), and Elixhauser comorbidity score.

For clarity, we present only two regression models (the two SSSI regression models) in the main results in Table 3 with the remaining models provided in the Supplementary Material (Table S2). The regression model for superficial surgical site infection (SSSI) using the F0.5 threshold did not yield any significant association between hospitals and PCs. This indicates no statistically significant variation in SSSI rates between hospitals under this threshold setting when compared to the reference hospital (hospital A).

**Table 3.** Regression Model results for Superficial Surgical Site Infection (SSSI) including Odds Ratios (OR), 95% Confidence Intervals (CI), and p-values for both F0.5 and F2 Thresholds. Statistically significant results are highlighted in bold. Regression Model results includes Odds Ratios (OR), 95% Confidence Intervals (CI), and p-values for both F0.5 and F2 Thresholds.

|  | SSSI (F0.5 thresholds) |  | SSSI (F2 thresholds) |  |
| --- | --- | --- | --- | --- |
|  | Odds Ratio (95% CI) | p-value | Odds Ratio (95% CI) | p-value |
| **Const** | **19.4 (8.09-46.6)** | **3.09e<sup>-11</sup>** | **3.61 (2.63-4.95)** | **2.30e<sup>-15</sup>** |
| **Hospital B** | 0.913 (0.265-3.14) | 0.885 | 1.49 (0.985-2.26) | 0.0589 |
| **Hospital C** | 1.41 (0.132-15.1) | 0.776 | **2.31 (1.04-5.15)** | **0.0401** |
| **Hospital D** | 0.919 (0.382-2.21) | 0.850 | **1.45 (1.08-1.93)** | **0.0121** |
| **Hospital E** | 1.00 (0.238-4.21) | 0.998 | 1.48 (0.913-2.39) | 0.112 |
| **Hospital F** | 0.993 (0.232-4.26) | 0.993 | 1.26 (0.823-1.94) | 0.286 |
| **Hospital G** | 0.578 (0.158-2.12) | 0.408 | **1.93 (1.16-3.24)** | **0.0121** |
| **Hospital H** | 0.981 (0.277-3.47) | 0.976 | **2.43 (1.51-3.91)** | **2.37e<sup>-4</sup>** |
| **Hospital I** | 0.901 (0.367-2.22) | 0.821 | **1.76 (1.27-2.42)** | **6.06e<sup>-4</sup>** |
| **Hospital J** | 0.964 (0.357-2.61) | 0.943 | **1.49 (1.04-2.13)** | **0.0282** |
| **Hospital K** | 0.831 (0.210-3.29) | 0.792 | 1.67 (0.940-2.95) | 0.0803 |
| **Hospital L** | 1.34 (0.301-5.92) | 0.703 | 1.62 (0.999-2.62) | 0.0504 |
| **Age** | 0.999 (0.984-1.02) | 0.949 | **0.986 (0.981- 0.991)** | **1.93e<sup>-7</sup>** |
| **Elixscore** | 0.985 (0.984-1.02) | 0.733 | 0.984 (0.957-1.01) | 0.261 |
| **Sex, M** | 0.987 (0.902-1.08) | 0.968 | 0.855 (0.693-1.06) | 0.145 |

However, when applying the F2 thresholds, six hospitals showed a statistically significant association with SSSI (p < 0.05). These hospitals (hospital C, D, G, H, I, and J) all had an increased odds ratio (OR) ranging from 1.45 to 2.43, indicating an increased risk of SSSI in these hospitals compared to the reference hospital (hospital A). Two hospitals (hospital B and L) exhibited a trend toward significance with p-values 0.0589 and 0.0504 respectively. Additionally, in this regression model (F2 thresholds), age was also statistically significant with an OR of 0.986 (95% CI: 0.981-0.991, p-value = 1.93e^-7^) suggesting a slight decrease in the risk of SSSI as age increases. All results for the two regression models on SSSI, including ORs, 95% confidence intervals, and p-values, are detailed in Table 3.

## DISCUSSION

The primary objective of this study was to assess the feasibility of an automated registry for postoperative complications (PCs) using NLP algorithms for quality surveillance and improvement. Our findings demonstrate that NLP can effectively detect PCs, potentially automating data curation for quality registries. However, despite this technological advancement, PC rates over time remained mostly stable or increased—both for individual PCs and overall hospital rates—suggesting limited improvement during the study period.

It is well documented that PCs are often underreported when monitored using administrative data sources [8,17–19]. While NLP algorithms offer a potential improvement in this monitoring process, their implementation requires determining a cutoff value at which the models flag a PC as present—that is, setting a threshold. This threshold directly affects the model’s performance metrics (sensitivity, specificity, positive predictive value, and negative predictive value). Therefore, it is crucial to balance these metrics when using NLP models in a clinical setting, as it is not possible to achieve perfect performance across all measures. To illustrate this trade off, we present our results using two different threshold sets. As illustrated in our findings, threshold selection significantly influenced the number of detected PCs, despite high NLP model ROC AUC scores. Sensitivity-optimized thresholds nearly doubled the total number of detected PCs compared to PPV-optimized thresholds (15,892 vs. 8,512), a pattern also reflected in individual PC rates. This resulted in a total PC rate ranging from 9.14–17.1%, aligning with findings from a systematic review and meta-analysis by Anderson et al., which reported an adverse event rate of 14.4%[20].

When comparing our results with existing literature, it is evident that countries with large-scale quality improvement initiatives, such as the ACS NSQIP and HRRP (Hospital Readmission Reduction Program), typically report declining PC rates over time[21–26]. The American College of Surgeons has documented continual improvements in surgical outcomes for participating hospitals[25], and Cohen et al. found that longer participation in the ACS NSQIP was associated with further reductions in adverse events[23].

While some studies align with our findings of stable PC rates[27,28], the broader literature suggests that reducing PCs requires active interventions rather than passive monitoring[29,30]. This aligns with the Salzburg Statement, which emphasizes the need for real-time data collection and reducing the burden of data analysis to improve patient safety[1]. NLP-driven quality monitoring directly addresses these principles by enhancing efficiency and facilitating faster identification of complications.

A critical finding of this study is that threshold selection influences not only PC incidence but also hospital benchmarking. While some models produced consistent findings across thresholds, others, such as the association between superficial surgical site infection (SSSI) and hospital location, varied substantially. PPV-optimized thresholds (F0.5) did not identify any significant hospital variation, whereas sensitivity-optimized thresholds (F2) flagged six hospitals with a significantly increased risk of SSSI. This underscores the importance of carefully considering threshold selection in NLP-driven quality monitoring, as it can directly impact hospital comparisons and quality improvement initiatives. Overlooking threshold effects may lead to misguided interventions or missed quality discrepancies.

The prevailing literature on AI-driven healthcare solutions emphasizes the potential of machine learning models, with many studies demonstrating performance metrics that match or exceed human curation[11,12,31–34]. Key advantages include reduced costs, efficient resource allocation, and faster data processing[11,12,35,36]. One of the key strengths of this study is its nuanced evaluation of NLP model performance beyond ROC AUC scores. By demonstrating the impact of threshold selection, this study highlights the limitations of NLP models in clinical settings while ensuring transparency. Additionally, this is the first study of its kind in a Danish setting and among the first internationally to explore the use of NLP for a comprehensive, automated PC registry designed for quality improvement.

However, despite these benefits, limitations to this study must be acknowledged. As illustrated, the ROC AUC metric might not fully reflect the performance of models in a real-world setting. ROC AUC primarily evaluates a model’s ability to distinguish between classes (e.g., presence or absence of a PC) but this can be misleading in imbalanced datasets. Nevertheless, upwards of 93% of studies in this field report ROC AUC as the primary performance metric[37]. While ROC AUC provides valuable insight, one could argue that additional performance metrics, including threshold selection strategies and precision-recall curves, should be considered before deploying NLP models in any clinical setting[37–39]. Moreover, the strong dependence on threshold selection means that the approach in this study is not ready for direct clinical implementation. Before NLP models can be used for automated PC registries, external validation across diverse populations and healthcare settings is necessary. Model performance varies across populations, posing risks of bias and reduced reliability.

While this study presents several measures for a possible future surgical quality register, we do not present an exhaustive list, and additional data on further surgical procedures, specialties, and hospitals would be valuable to include. Additionally, we did not account for changes in the proportion of high-risk surgeries within the included procedures over time.

Furthermore, while the NLP models offer efficiency, they are not without risk. Compared to human curation, which remains the gold standard in surgical quality monitoring, NLP models are susceptible to false positives and false negatives, potentially impacting clinical decision-making. Lastly, the use of AI in healthcare also raises ethical, transparency, and patient safety concerns.

In conclusion, this study illustrates how AI can be integrated into traditionally resource-intensive quality improvement projects. Our findings highlight the importance of considering multiple performance metrics beyond ROC AUC to ensure the reliable and effective use of NLP models. Future research should focus on exploring threshold selection, validating models in diverse settings, and addressing the practical challenges of AI implementation in clinical care.

## Submission category

Original article

## Data availability statement

In compliance with GDPR-regulations, the authors are unable to share the data. Access to the dataset can be permitted through the Danish Patients Safety Board (Styrelsen for Patientsikkerhed) and the Danish Capital Region Data Safety Board (Videnscenter for Dataanmeldelser).

## Author contributions (CRediT)

EED, MS, AB, ASM, JWJ, and AT conceptualized the study and designed the methodology. EED, ASM and AB performed data curation and formal analysis. EED compiled relevant data and information, drafted the manuscript, and created visual elements. MS supervised the study while MS, ASM, AB, JWJ, and AT reviewed and revised the manuscript.

