## Supplementary data for "TOWARDS AN AI-DRIVEN REGISTRY FOR POSTOPERATIVE COMPLICATIONS: A PROOF-OF-CONCEPT STUDY EVALUATING THE OPPORTUNITIES AND CHALLENGES OF AI-MODELS"

**Supplementary Material**

**Table S1**

The American College of Surgeons NSQIP definitions of the studied postoperative complications. ACS NSQIP® OPERATIONS MANUAL, Chapter 4, June 2018

| **Pneumonia** | |
| --- | --- |
| **DEFINITION**: | Pneumonia is an infection of one or both lungs caused by bacteria, viruses, fungi, or aspiration. Pneumonia can be community acquired or acquired in a healthcare setting. |
| **CRITERIA**: | The case must meet Radiology (A) criteria **AND** ONE of the following TWO Signs/Symptoms/Laboratory (B) scenarios as listed below within the 30 days after the primary procedure.  **A. Radiology:**  **ONE** chest radiological exam (X-ray or CT)* demonstrating at least **ONE** of the following:   - Infiltrate - Consolidation - Opacity - Cavitation   **OR**   - A diagnosis of pneumonia is rendered by an attending physician based on the findings demonstrated on a chest radiological exam (X-ray or CT).   *Two imaging tests are required for patients with underlying pulmonary or cardiac disease (e.g., respiratory distress syndrome, bronchopulmonary dysplasia, pulmonary edema, or chronic obstructive pulmonary disease).  **B.** **Signs/Symptoms/Laboratory:**  SCENARIO #1  At least **ONE** of the following:   - Fever (>38 ֯C or >100.4 ֯F) with no other recognized cause - Leukopenia (<4000 WBC/mm^3^) or leukocytosis (≥12,000 WBC/ mm^3^) - For adults ≥ 70 years old, altered mental status with no other recognized cause   **AND**  At least **ONE** of the following:   - 5% Bronchoalveolar lavage (BAL) -obtained cells contain intracellular bacteria on direct microscopic exam (e.g., Gram stain) - Positive growth in blood culture not related to another source of infection - Positive growth in culture of pleural fluid - Positive quantitative culture from minimally contaminated lower respiratory tract (LRT) specimen (e.g., BAL or protected specimen brushing)   **OR**  SCENARIO #2  At least **ONE** of the following:   - Fever (>38 ֯C or >100.4 ֯F) with no other recognized cause - Leukopenia (<4000 WBC/ mm^3^) or leukocytosis (≥12,000 WBC/ mm^3^) - For adults ≥ 70 years old, altered mental status with no other recognized cause   **AND**  At least **TWO** of the following:   - New onset of purulent sputum, or change in character of sputum, or increased respiratory secretions, or increased suctioning requirements - New or worsening cough, dyspnea, or tachypnea - Rales (crackles) or rhonchi - Worsening gas exchange (e.g., O2 desaturations (e.g., PaO2/FiO2 ≤ 240), increased oxygen requirements, or increased ventilator demand) |
| **SCENARIOS**  **TO ASSIGN:** | - When pneumonia criteria are met, and the patient has aspiration pneumonitis or aspiration pneumonia - Clearly defined description of airspace disease (such as “suspicious for aspiration or infection”) would meet radiologic criteria. |
| **SCENARIOS TO NOT ASSIGN:** | - Documentation of undefined airspace disease and densities on an X-ray would not qualify. Airspace disease may be referring to infection. If this is not clearly defined by the radiologist this description cannot be used. Densities may be referring to tumors rather than evidence of infection. - A sputum culture is not considered a lower respiratory tract (LRT) specimen and cannot be utilized to assign Pneumonia. - Pneumonia progressing to another lobe is not a new Pneumonia. - At the time of care, the physician documents that the patient does not have pneumonia and treats the patient for a noninfectious pulmonary process (e.g.,pulmonary edema, pleural effusion, pulmonary hemorrhage, atelectasis) and subsequent serial radiographs demonstrate a rapid decrease or resolution in the positive radiological findings. |

| **Urinary tract infection** | |
| --- | --- |
| **DEFINITION**: | An infection in the urinary tract (kidneys, ureters, bladder, and urethra). |
| **CRITERIA:** | Must be noted within 30 days after the primary procedure **AND** patient must meet **ONE** of the following A **OR** B below:  **A: ONE** of the following six criteria:   - fever (>38 ֯C or >100.4 ֯F) - urgency - frequency - dysuria - suprapubic tenderness - costovertebral angle pain or tenderness   **AND**   1. A urine culture of > 100,000 colonies/ml urine with no more than two species of organisms. Signs and symptoms should be reported within 72 hours prior to a urine culture being sent or 24 hours after the culture was sent.   **OR**  **B: TWO** of the following six criteria:   2. fever (>38 ֯C or >100.4 ֯F) 3. urgency 4. frequency 5. dysuria 6. suprapubic tenderness 7. costovertebral angle pain or tenderness   **AND**  **At least one** of the following:   - Two urine cultures with repeated isolation of the same uropathogen with >100 colonies/ml urine in non-voided specimen. Signs and symptoms should be reported within 72 hours prior to a urine culture being sent or 24 hours after the culture was sent. - Urine culture with < 100,000 colonies/ml urine of single uropathogen in patient being treated with appropriate antimicrobial therapy. Signs and symptoms should be reported within 72 hours prior to a urine culture being sent or 24 hours after the culture was sent. - Physician or Advanced Practitioner diagnosis - Physician or Advanced Practitioner institutes appropriate antimicrobial therapy |
| **SCENARIOS TO NOT ASSIGN:** | - Asymptomatic UTI which are treated or untreated - Patients with Foley catheters who do not display signs or symptoms |

| **Sepsis** | |
| --- | --- |
| **DEFINITION**: | Sepsis is the systemic response to infection. |
| **CRITERIA**: | Report the **most significant** level using criteria below: Septic Shock is more severe than sepsis. Criteria must be noted within 30 days after the primary procedure**.**  **A. Five Clinical Signs of SIRS (need two):**   1. Temp >38 ֯C or >100.4 ֯F or < 36 ֯C (96.8 ֯F) 2. HR >90 bpm 3. RR >20 breaths/min or PaCO_2_ <32 mmHg (<4.3 kPa) 4. WBC >12,000 cell/mm^3^, <4000 cells/ mm^3^, or >10% immature (band) forms 5. Anion gap acidosis: this is defined by either: (check with your Lab on calculation)   a. [Na + K] – [Cl + HCO_3_ (or serum CO_2_)]. If >16, then an anion gap acidosis is present  b. Na – [Cl + HCO_3_ (or serum CO_2_)]. If > than 12, then an anion gap acidosis is present  **AND**  **B. Sepsis: Either scenario 1, 2, 3, or 4:**  **Scenario 1: Following the primary procedure or a reoperation.**  The patient must meet SIRS criteria (any two of the five) **and ONE** of the following:   a. Positive blood culture b. Clinical documentation of purulence or positive culture from any site for which there is correlating physician documentation that the site was thought to be the **acute** cause of the septic picture   **OR**  **Scenario 2: Immediately following and up to 8 hours after the primary procedure or a reoperation**  Immediately following and up to 8 hours after the primary procedure or reoperation an elevated heart rate and elevated respiratory rate together will **not** satisfy SIRS criteria. SIRS criteria must be met utilizing at least one criterion of temperature, leukocyte count, or anion gap acidosis if an elevated heart rate or elevated respirations is used.  The patient must meet SIRS criteria as specified above **AND** one of the following findings during the primary procedure or reoperation:   a. Confirmed ischemic/infarcted bowel (for instance requiring resection) b. Purulence in the operative site c. Enteric/gastric contents in the operative site d. Positive intraoperative culture   **OR**  **Scenario 3: 8- 48 hours after the Primary procedure**  The patient must meet SIRS criteria (any two of the five) between 8 and 48 hours after the principal operative procedure **AND** one of the following findings during the primary procedure:   a. Confirmed ischemic/infarcted bowel (for instance requiring resection) b. Purulence in the operative site c. Enteric/gastric contents in the operative site d. Positive intraoperative culture   **OR**  **Scenario 4**: **48 hours before or 8 – 48 hours after a Reoperation**  The patient must meet SIRS criteria (any two of the five) within 48 hours before or between 8 and 48 hours after a subsequent reoperation **AND** one of the following findings during a subsequent operation:   a. Confirmed ischemic/infarcted bowel (for instance requiring resection) b. Purulence in the operative site c. Enteric/gastric contents in the operative site d. Positive intraoperative culture |
| **SCENARIOS TO ASSIGN:** | - The presence of pneumatosis along with the presence of SIRS. |
| **SCENARIOS TO NOT ASSIGN:** | - Patient meets SIRS criteria; however, only a positive culture from a chronic leg wound, which is unchanged in its chronic appearance. - In cases with documented explanation or evidence that the criteria being reviewed for SIRS (i.e. HR, RR, Temp) are likely due to a cause other than an inflammatory or infectious process. |

| **Septic shock** | |
| --- | --- |
| **DEFINITION**: | Sepsis is considered severe when it is associated with organ and/or circulatory dysfunction. |
| **CRITERIA**: | Criteria must be noted within 30 days after the primary procedure.  Report variable if patient meets SIRS criteria (A) AND meets the criteria of Septic Shock below:   1. Sepsis criteria   **AND**   2. Has documented organ and/or circulatory dysfunction.    - Examples of organ dysfunction: oliguria, acute alteration in mental status, acute respiratory distress.    - Examples of circulatory dysfunction: hypotension requiring inotropic or vasopressor agents. |
| **SCENARIOS TO NOT ASSIGN:** | - Cardiogenic, neurogenic, distributive or hypovolemic shock, in the absence of meeting above criteria |

| **Superficial incisional surgical site infection** | |
| --- | --- |
| **DEFINITION**: | Superficial Incisional SSI is an infection that involves only skin or subcutaneous tissue of the surgical incision. |
| **CRITERIA**: | An infection that occurs within 30 days after the primary procedure AND the infection involves only skin or subcutaneous tissue of the incision AND at least ONE of the following:  A. Purulent drainage, with or without laboratory confirmation, from the superficial incision  B. Organisms isolated from an aseptically obtained culture of fluid or tissue from the superficial incision  C. Superficial incision is deliberately opened by the surgeon (see note below)  **AND**  At least one of the following signs or symptoms of infection:   - pain or tenderness - localized swelling - redness - heat   If the patient meets criterion C and the surgical incision is cultured, a negative culture result would exclude the assignment of Superficial SSI based on criterion C only.  NOTE: Please also refer to Appendix H. Superficial Incisional SSI Algorithm posted to the Resource Portal,Program Resources Tab, ACS NSQIP Operations Manual, Appendix H for additional guidance in assigning a Superficial SSI to a case utilizing criterion C.  D. Diagnosis of superficial incisional SSI by the surgeon or attending physician |
| **SCENARIOS TO ASSIGN:** | - Superficial SSI which occurs at a drain site, in which the drain was placed during the primary procedure. - If patient meets criteria A or D, a negative culture of the surgical incision will not affect the assignment of this occurrence |
| **SCENARIOS TO NOT ASSIGN:** | - Stitch abscess (minimal inflammation and discharge confined to the points of suture penetration) - Infected burn wound - Incisional SSI that extends into the fascia and muscle layers (see Deep Incisional SSI) - If wound closure for the primary procedure is documented as “superficial layers are left open” or “no layers closed” - Report infection that involves both superficial and deep incision sites as Deep Incisional SSI - Diagnosis of cellulitis would not meet criterion D - Diagnosis of superficial vaginitis, after vaginal surgery would not meet criterion D - • Diagnosis of superficial oral thrush after oral surgery would not meet criterion D |

| **Unplanned readmission (30 day readmission)** | |
| --- | --- |
| **DEFINITION**: | Patients who were discharged from their index hospital stay or encounter (whether inpatient or outpatient basis) after their primary procedure, and within 30 days of the primary procedure, are subsequently formally readmitted by a qualified practitioner as an inpatient to an acute care bed  OR  Otherwise have a subsequent hospital (or facility-based) encounter (receiving outpatient or observation services) that crosses at least two midnights. |
| **CRITERIA**: | Enter each hospital inpatient readmission separately.  Report any readmission (to the same or another hospital), for any reason, within 30 days after the primary procedure. The readmission has to be classified as an “inpatient” stay by the readmitting hospital or reported by the patient/family as such.  For each readmission enter the following:  Date of Readmission, if known, or select “Unknown”  Information Source:   - Medical record - Patient/Family report - Other   Unplanned Readmission at the time of the primary procedure?   - Yes - No   Readmission Related to Principal Procedure?  *“Yes” is the default answer unless it is definitively indicated that the readmission is not related to the primary procedure.*   - Yes - No   Primary Suspected Reason for Readmission:  Choose the primary suspected reason (from the list below) or “Other” and then enter the ICD code, or if code unknown please describe the reason for the readmission. Choosing one of these occurrences does not indicate that the NSQIP criteria for the occurrence were met; it merely indicates that this diagnosis was given as a reason for readmission.   - Superficial Incisional SSI - Deep Incisional SSI - Organ/Space SSI - Wound Disruption - Pneumonia - Unplanned Intubation - Pulmonary Embolism - On Ventilator > 48 Hours - Progressive Renal Insufficiency - Acute Renal Failure - Urinary Tract Infection (UTI) - Stroke/CVA - Cardiac Arrest Requiring CPR - Myocardial Infarction - Transfusion Intra/Postop (RBC within the First 72 Hrs of Surgery Start Time) - Vein Thrombosis Requiring Therapy - Clostridium Difficile (C.diff) Colitis - Sepsis - Septic Shock - Other |
| **SCENARIOS TO ASSIGN:** | - A patient returns to the Emergency Department at your site and is admitted to an acute care bed which includes outpatient or observation services. Utilize the date the order is written for the transfer to the acute care bed as the start date for the hospital stay or encounter when assessing for readmission. - If a patient is admitted to an outside Emergency Department and you do not have access to this site’s orders, utilize the patient arrival date to that outside Emergency Department as the hospital stay or encounter start date when assessing for readmission. |
| **SCENARIOS TO NOT ASSIGN:** | - If a patient is transferred from your acute care site directly to another acute care site, this is not a readmission. An example of this would be a transfer to a higher level of care or for a procedure that cannot be done at your institution. |

| **Unplanned reoperation (30-Day Unplanned Return to OR)** | |
| --- | --- |
| **DEFINITION**: | A return to the OR that was not planned at the time of the primary procedure. |
| **CRITERIA**: | Enter each unplanned reoperation separately.  Report any reoperation, for any reason, within 30 days after the primary procedure which was not planned prior to or at the time of the primary procedure.  **For each unplanned return to OR enter the following:**   - Date of Unplanned Return to OR, if known, or select “Unknown” - Unplanned Return to OR Procedure(s)   Enter the CPT® code(s) which corresponds to the procedures performed during the return to the OR. If no CPT® code is available, enter a description in Procedure Notes.   - Unplanned Return to OR Related to Primary Procedure?   “Yes” is the default answer unless it is definitively indicated that the unplanned return to the OR is not related to the Primary Procedure.   - Yes - No (If related to the Primary Procedure, record the ICD 10 Code, or provide a Diagnosis Description if code is not documented or is unavailable) |
| **SCENARIOS TO ASSIGN:** | - If the Primary Procedure was performed in the main OR (regardless of the specific type of suite, such as vascular, cardiac, gyne) |
| **SCENARIOS TO NOT ASSIGN:** | Scenarios to Clarify (Do Not Assign Variable):  If there is documentation preoperatively or intraoperatively that the patient requires a return to the OR following the Primary Procedure  A return to the OR if:   - An unintended principal procedure is aborted due to patient physiology and is rescheduled for the completion of the initial procedure at a later date. - Unanticipated findings are discovered, such as a progressed disease state, during the Primary Procedure requiring additional or subsequent operations. - An unintentional bowel perforation occurred during the Primary Procedure and intraoperatively was noted will require additional procedures or closure at a later date.   This definition is not meant to capture patients who go back to the operating room within 30 days for a follow-up procedure based on the pathology results from the primary procedure or concurrent procedure |

**Table S2** - Regression Model results for remaining postoperative complications: pneumonia, urinary tract infection (UTI), sepsis, septic shock, unplanned readmission, and unplanned reoperation. Results include Odds Ratios (OR), 95% Confidence Intervals (CI), and p-values for both F0.5 and F2 thresholds.

|  | **Pneumonia (F0.5 thresholds)** | | **Pneumonia (F2 thresholds)** | |
| --- | --- | --- | --- | --- |
|  | **Odds Ratio (95% CI)** | **p-value** | **Odds Ratio (95% CI)** | **p-value** |
| **Const** | 0.914 (0.294-2.84) | 0.876 | **0.424 (0.210-0.857)** | **0.0168** |
| **Hospital B** | 0.921 (0.386-2.20) | 0.854 | 0.891 (0.491-1.62) | 0.704 |
| **Hospital C** | 0.824 (0.131- 5.17) | 0.836 | 0.830 (0.220-3.13) | 0.783 |
| **Hospital D** | 1.00 (0.505-1.99) | 0.998 | 0.903 (0.577-1.41) | 0.657 |
| **Hospital E** | 0.920 (0.300-2.82) | 0.884 | 0.827 (0.397-1.72) | 0.611 |
| **Hospital F** | 1.03 (0.458-2.31) | 0.946 | 1.03 (0.583-1.80) | 0.931 |
| **Hospital G** | 0.859 (0.222-3.32) | 0.826 | 0.903 (0.359-2.28) | 0.829 |
| **Hospital H** | 1.00 (0.260-3.86) | 0.998 | 0.931 (0.383-2.27) | 0.875 |
| **Hospital I** | 0.997 (0.439-2.27) | 0.994 | 0.888 (0.515-1.53) | 0.669 |
| **Hospital J** | 0.951 (0.292-3.10) | 0.933 | 1.04 (0.470-2.31) | 0.920 |
| **Hospital K** | 0.805 (0.163-3.97) | 0.790 | 0.804 (0.302-2.14) | 0.663 |
| **Hospital L** | 1.12 (0.435-2.86) | 0.816 | 0.930 (0.498-1.74) | 0.820 |
| **Age** | 1.00 (0.986-1.02) | 0.875 | 1.00 (0.994-1.01) | 0.499 |
| **Elixscore** | 1.00 (0.956-1.05) | 0.879 | 0.996 (0.967-1.03) | 0.814 |
| **Sex, M** | 1.03 (0.663-1.59) | 0.907 | 1.04 (0.781-1.39) | 0.778 |

|  | **UTI (F0.5 thresholds)** | | **UTI (F2 thresholds)** | |
| --- | --- | --- | --- | --- |
|  | **Odds Ratio (95% CI)** | **p-value** | **Odds Ratio (95% CI)** | **p-value** |
| **Const** | 2.18 (0.838-5.70) | 0.110 | **0.340 (0.237- 0.488)** | **5.20e^-9^** |
| **Hospital B** | 1.25 (0.203-7.67) | 0.811 | 0.973 (0.510-1.86) | 0.935 |
| **Hospital C** | 1.41 (0.0597-33.5) | 0.830 | 1.07 (0.395-2.88) | 0.899 |
| **Hospital D** | 1.09 (0.422-2.81) | 0.859 | 1.16 (0.828-1.63) | 0.384 |
| **Hospital E** | 0.904 (0.174-4.71) | 0.905 | 1.01 (0.543-1.87) | 0.980 |
| **Hospital F** | 1.19 (0.0913-15.5) | 0.894 | 0.879 (0.408-1.90) | 0.743 |
| **Hospital G** | 1.14 (0.169-7.65) | 0.894 | 0.930 (0.492-1.76) | 0.824 |
| **Hospital H** | 1.04 (0.158-6.77) | 0.971 | 1.04 (0.471-2.28) | 0.928 |
| **Hospital I** | 1.05 (0.330-3.36) | 0.930 | 1.11 (0.713-1.74) | 0.635 |
| **Hospital J** | 1.05 (0.374-2.95) | 0.925 | 1.14 (0.783-1.67) | 0.487 |
| **Hospital K** | 1.07 (0.323-3.56) | 0.909 | 1.04 (0.677-1.60) | 0.853 |
| **Hospital L** | 1.01 (0.202-5.05) | 0.990 | 0.950 (0.524-1.72) | 0.867 |
| **Age** | 0.997 (0.983-1.01) | 0.630 | 1.00 (0.994-1.01) | 0.919 |
| **Elixscore** | 1.01 (0.941-1.07) | 0.869 | 1.00 (0.977-1.03) | 0.895 |
| **Sex, M** | 0.937 (0.543-1.62) | 0.814 | 0.944 (0.759-1.17) | 0.604 |

|  | **Sepsis (F0.5 thresholds)** | | **Sepsis (F2 thresholds)** | |
| --- | --- | --- | --- | --- |
|  | **Odds Ratio (95% CI)** | **p-value** | **Odds Ratio (95% CI)** | **p-value** |
| **Const** | 1.233 (0.473-3.21) | 0.669 | **0.445 (0.239-0.830)** | **0.0108** |
| **Hospital B** | 1.04 (0.421-2.59) | 0.925 | 0.903 (0.479-1.70) | 0.752 |
| **Hospital C** | 1.83 (0.317-10.6) | 0.499 | 1.43 (0.432-4.72) | 0.559 |
| **Hospital D** | 0.810 (0.410-1.60) | 0.546 | 0.732 (0.451-1.19) | 0.205 |
| **Hospital E** | 0.682 (0.211-2.20) | 0.521 | 0.683 (0.302-1.54) | 0.360 |
| **Hospital F** | 0.667 (0.181-2.46) | 0.543 | 0.512 (0.234-1.12) | 0.0938 |
| **Hospital G** | 0.784 (0.0456- 13.5) | 0.867 | 0.541 (0.156-1.87) | 0.332 |
| **Hospital H** | 0.930 (0.0161-53.7) | 0.972 | 0.404 (0.0785-2.08) | 0.278 |
| **Hospital I** | 0.870 (0.347-2.18) | 0.767 | 0.710 (0.384-1.31) | 0.274 |
| **Hospital J** | 1.31 (0.476-3.62) | 0.597 | 1.09 (0.537-2.20) | 0.814 |
| **Hospital K** | 0.694 (0.155-3.11) | 0.633 | **0.413 (0.174-0.981)** | **0.0450** |
| **Hospital L** | 0.925 (0.238-3.59) | 0.910 | 0.581 (0.270-1.25) | 0.165 |
| **Age** | 1.00 (0.988-1.02) | 0.774 | 1.01 (0.996-1.01) | 0.239 |
| **Elixscore** | 0.998(0.947-1.05) | 0.953 | 0.999 (0.966-1.03) | 0.977 |
| **Sex, M** | 0.991 (0.597-1.64) | 0.972 | 1.15 (0.824-1.60) | 0.417 |

|  | **Septic shock (F0.5 thresholds)** | | **Septic shock (F2 thresholds)** | |
| --- | --- | --- | --- | --- |
|  | **Odds Ratio (95% CI)** | **p-value** | **Odds Ratio (95% CI)** | **p-value** |
| **Const** | 2.19 (0.0176-271) | 0.750 | 0.289 (0.0537-1.56) | 0.148 |
| **Hospital B** | 0.700 (0.0636-7.70) | 0.771 | 1.34 (0.382-4.71) | 0.648 |
| **Hospital C** | 0.861 (0.0350-21.2) | 0.927 | 1.73 (0.148-20.2) | 0.663 |
| **Hospital D** | 0.862 (0.0808-9.19) | 0.902 | 0.791 (0.277-2.26) | 0.661 |
| **Hospital E** | 0.595 (0.0223-15.9) | 0.757 | 0.740 (0.198-2.77) | 0.655 |
| **Hospital F** | 0.791 (0.103-6.08) | 0.822 | 1.10 (0.362- 3.36) | 0.863 |
| **Hospital G** | NA | NA | 0.683 (0.0375-12.5) | 0.797 |
| **Hospital H** | NA | NA | 0.754 (0.0425-13.4) | 0.848 |
| **Hospital I** | 1.01 (0.0755-13.6) | 0.993 | 1.01 (0.302-3.39) | 0.985 |
| **Hospital J** | 0.880 (0.0107-72.4) | 0.955 | 0.850 (0.104-6.96) | 0.880 |
| **Hospital K** | NA | NA | 0.715 (0.0808-6.33) | 0.763 |
| **Hospital L** | 0.748 (0.100-5.57) | 0.777 | 0.915 (0.307-2.73) | 0.873 |
| **Age** | 0.992 (0.930-1.06) | 0.815 | 1.00 (0.981-1.03) | 0.744 |
| **Elixscore** | 1.03 (0.884-1.19) | 0.741 | 0.997 (0.942-1.05) | 0.907 |
| **Sex, M** | 1.07 (0.277-4.11) | 0.926 | 1.05 (0.563-1.94) | 0.889 |

|  | **Unplanned Readmission (F0.5 thresholds)** | | **Unplanned Readmission (F2 thresholds)** | |
| --- | --- | --- | --- | --- |
|  | **Odds Ratio (95% CI)** | **p-value** | **Odds Ratio (95% CI)** | **p-value** |
| **Const** | **10.3 (6.85-15.5)** | **4.23e^-29^** | **6.91 (5.03-9.50)** | **8.74e^-33^** |
| **Hospital B** | 0.857 (0.509-1.44) | 0.560 | 0.906 (0.596-1.38) | 0.646 |
| **Hospital C** | 0.838 (0.364-1.93) | 0.677 | 0.941 (0.469-1.89) | 0.863 |
| **Hospital D** | 0.920 (0.633-1.34) | 0.662 | 0.835 (0.624-1.12) | 0.222 |
| **Hospital E** | 1.13 (0.615-2.06) | 0.699 | 1.08 (0.673-1.72) | 0.759 |
| **Hospital F** | 1.27 (0.708-2.28) | 0.422 | 1.41 (0.870-2.29) | 0.163 |
| **Hospital G** | 1.02 (0.543-1.90) | 0.959 | 1.06 (0.641-1.74) | 0.830 |
| **Hospital H** | 0.959 (0.539-1.71) | 0.886 | 1.13 (0.694-1.82) | 0.633 |
| **Hospital I** | 0.943 (0.599-1.49) | 0.801 | 1.01 (0.704-1.46) | 0.942 |
| **Hospital J** | 0.704 (0.456-1.09) | 0.113 | **0.525 (0.385-0.717)** | **5.02e^-5^** |
| **Hospital K** | 0.842 (0.490-1.45) | 0.533 | **0.665 (0.451-0.981)** | **0.0398** |
| **Hospital L** | 1.12 (0.623-1.99) | 0.714 | 1.26 (0.775-2.04) | 0.354 |
| **Age** | 1.00 (0.995-1.01) | 0.836 | 1.00 (0.995-1.00) | 0.997 |
| **Elixscore** | 0.997 (0.968-1.03) | 0.812 | 1.01 (0.987-1.03) | 0.427 |
| **Sex, M** | 0.971 (0.766-1.23) | 0.807 | 0.945 (0.788-1.13) | 0.546 |

|  | **Unplanned Reoperation (F0.5 thresholds)** | | **Unplanned Reoperation (F2 thresholds)** | |
| --- | --- | --- | --- | --- |
|  | **Odds Ratio (95% CI)** | **p-value** | **Odds Ratio (95% CI)** | **p-value** |
| **Const** | **228.5 (28.46-1834)** | **3.21e^-7^** | **23.0 (15.4-34.3)** | **2.09e^-53^** |
| **Hospital B** | 1.11 (0.0843-14.5) | 0.939 | 0.741 (0.485-1.13) | 0.163 |
| **Hospital C** | 1.08 (0.00880-134) | 0.974 | 1.05 (0.451-2.42) | 0.918 |
| **Hospital D** | 0.912 (0.146-5.68) | 0.921 | **0.596 (0.433-0.822)** | **1.57e^-3^** |
| **Hospital E** | 1.04 (0.0723-14.9) | 0.978 | 1.34 (0.773-2.34) | 0.295 |
| **Hospital F** | 1.28 (0.0950-17.1) | 0.850 | 1.10 (0.687-1.76) | 0.693 |
| **Hospital G** | 1.62 (0.0448-58.3) | 0.793 | 1.46 (0.795-2.66) | 0.224 |
| **Hospital H** | 1.36 (0.0634-29.3) | 0.843 | 1.33 (0.743-2.38) | 0.337 |
| **Hospital I** | 0.944 (0.125-7.11) | 0.955 | 1.09 (0.726-1.63) | 0.681 |
| **Hospital J** | 0.982 (0.0733-13.2) | 0.989 | **0.372 (0.255-0.544)** | **3.30e^-7^** |
| **Hospital K** | 0.970 (0.0256-36.7) | 0.987 | **0.327 (0.212-0.503)** | **3.73e^-7^** |
| **Hospital L** | 0.988 (0.0900-10.8) | 0.991 | 1.08 (0.674-1.74) | 0.740 |
| **Age** | 0.992 (0.962-1.02) | 0.630 | **0.980 (0.974-0.985)** | **1.67e^-13^** |
| **Elixscore** | 0.997 (0.853-1.17) | 0.973 | 1.02 (0.989-1.04) | 0.250 |
| **Sex, M** | 0.869 (0.261-2.89) | 0.819 | **0.751 (0.619- 0.911)** | **3.64e^-3^** |
